# Machine learning approach for ambient-light-corrected parameters and the Pupil Reactivity (PuRe) Score in smartphone-based pupillometry

**DOI:** 10.1101/2023.12.29.23300638

**Authors:** Aleksander Bogucki, Ivo A. John, Łukasz Zinkiewicz, Michał Jachura, Damian Jaworski, Hugo Chróst, Michal Wlodarski, Jakub Kałużny, Doug Campbell, Paul Bakken, Shawna Pandya, Radosław Chrapkiewicz, Sanjay G. Manohar

## Abstract

The pupillary light reflex (PLR) is the constriction of the pupil in response to light. The PLR in response to a pulse of light follows a complex waveform that can be characterized by several parameters. It is a sensitive marker of acute neurological deterioration, but is also sensitive to the background illumination in the environment in which it is measured. To detect a pathological change in the PLR, it is therefore necessary to separate the contributions of neuro-ophthalmic factors from ambient illumination. Illumination varies over several orders of magnitude and is difficult to control due to diurnal, seasonal, and location variations. We assessed the sensitivity of seven PLR parameters to differences in ambient light, using a smartphone-based pupillometer (AI Pupillometer, Solvemed Inc.). Nine subjects underwent 345 measurements in ambient conditions ranging from complete darkness (<5 lx) to bright lighting (≲10000 lx). Lighting most strongly affected the initial pupil size, constriction amplitude, and velocity. Nonlinear models were fitted to find the correction function that maximally stabilized PLR parameters across different ambient light levels. Next, we demonstrated that the lighting-corrected parameters still discriminated reactive from unreactive pupils. Ten patients underwent PLR testing in an ophthalmology outpatient clinic setting following the administration of tropicamide eye drops, which rendered the pupils unreactive. The parameters corrected for lighting were combined as predictors in a machine learning model to produce a scalar value, the Pupil Reactivity (PuRe) score, which quantifies pupil reactivity on a scale 0-5 (0, non-reactive pupil; 0-3, abnormal/”sluggish” response; 3-5, normal/brisk response). The score discriminated unreactive pupils with 100% accuracy and was stable under changes in ambient illumination across four orders of magnitude. This is the first time that a correction method has been proposed to effectively mitigate the confounding influence of ambient light on PLR measurements, which could improve the reliability of pupillometric parameters both in pre-hospital and inpatient care settings. In particular, the PuRe score offers a robust measure of pupil reactivity directly applicable to clinical practice. Importantly, the formulae behind the score are openly available for the benefit of the clinical research community.

## INTRODUCTION

The pupillary light reflex (PLR) is mediated by a well-understood neural pathway consisting of the retina, optic nerves, midbrain, brainstem, sympathetic and parasympathetic nervous systems ([12, 15, 20]). Consequently, the PLR is sensitive to a wide array of biological and environmental conditions ([9, 10, 14, 24, 32]), in particular ambient lighting. In order to elicit the PLR response, a single pulse of light is delivered to one eye, leading to pupil constriction (Fig. 1A). The dynamics of constriction can be characterized by several parameters. These are often measured as the initial pupil size, minimum pupil size, final pupil size, the size difference from initial to the minimum (constriction amplitude), the maximum constriction velocity, peak dilation velocity, the time to begin constriction (latency), and time to re-dilate (recovery time) (Fig. 1A) ([1, 19, 31]).

**FIG. 1:**
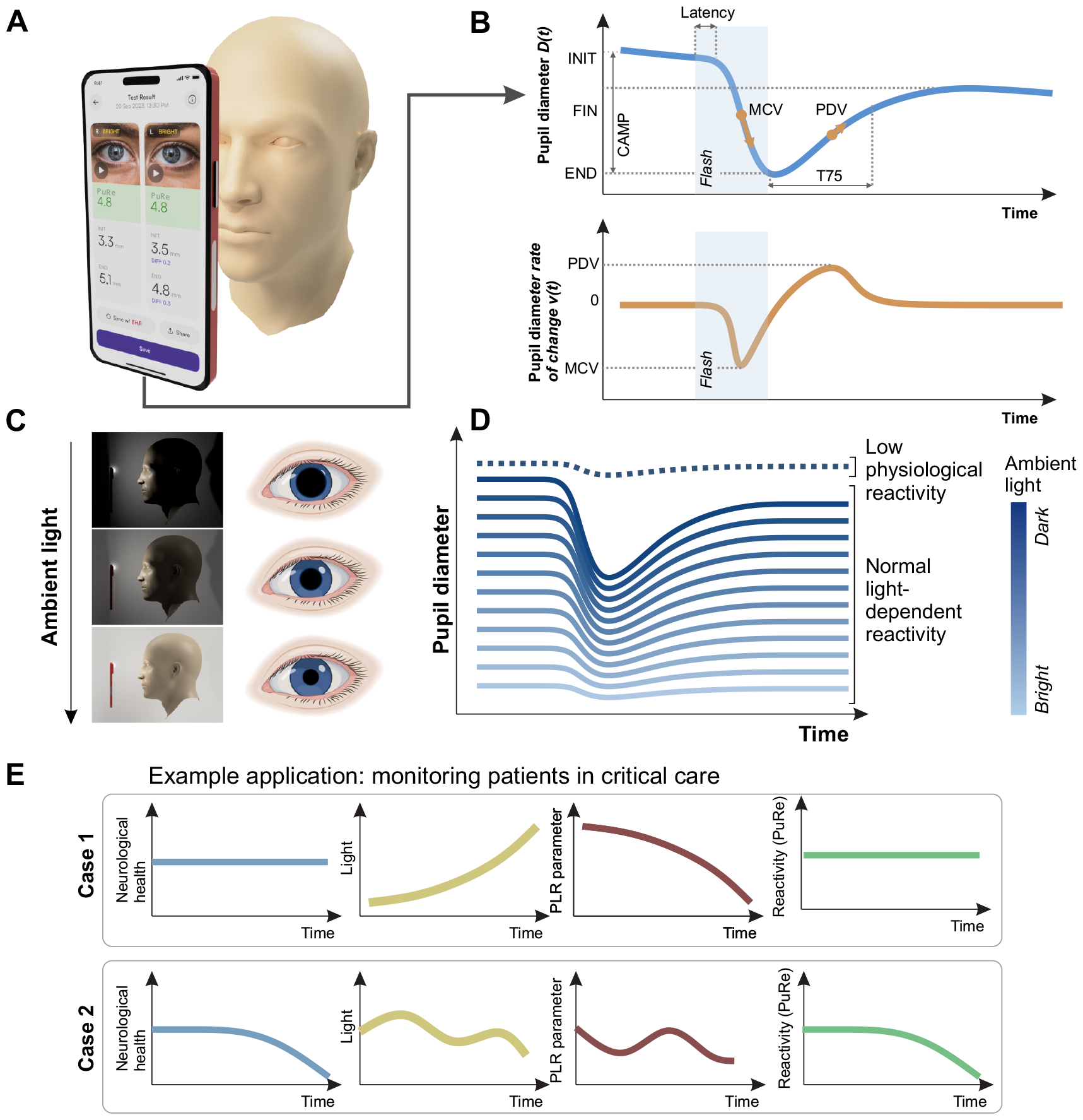
Establishing invariance to illumination in the pupillary light reflex (PLR) **(A)** We recorded the PLR using a Smartphone-based pupillometer. **(B)** The recording yields pupil diameter as a function of time. The curve can be characterised by 8 parameters (see Table I for descriptions). Shaded rectangle: flash stimulus window. Below: the rate of change of pupil diameter is used to estimate the velocity parameters. **(C)** Ambient light conditions significantly alter the baseline pupil diameter. This impacts the dynamics of the pupillary response to light **(D)** Under bright conditions, the response amplitude is attenuated (lower, pale blue lines), which could potentially be confused with an unreactive pupil (dotted line). **(E)** Top row: Ambient light variations, like those occurring throughout the day, can alter pupil parameters even without neurological deterioration, leading to potential misinterpretation. The Pupil Reactivity (PuRe) score is designed to monitor neurological health changes, while remaining stable despite varying light conditions. Bottom row: However, PuRe score should accurately reflect pupillary changes due to neurological health, even under variable lighting.

The PLR is an important marker of neurological decline in patients with conditions such as traumatic brain injury, stroke, infection, tumors, and aneurysms due to raised intracranial pressure (ICP) ([4–7, 9, 16, 18, 21, 25, 26, 28, 29, 34] Furthermore, the degree of PLR reactivity has recently been shown to hold prognostic value in patients with intracranial haemorrhage and traumatic brain injury ([4, 7, 21, 25, 26, 29]). It is therefore crucial for measured pupil parameters to be accurate. Current clinically used methods of assessing pupil size and reactivity do not control for the effects of ambient light, contributing to the lack of confidence in the test results including among trained professionals ([22]). Achieving inter-examiner reliability in pupil examinations performed with medical pentorches (penlights) proves challenging to practitioners ([23]), with the measurement reliability affected by varying levels of ambient light among other factors. Crucially, infra-red pupillometer devices can exhibit clinically relevant lighting dependence, with significant confounding effects reported previously in critical care patients ([10, 24]) calling for standardization of measurement conditions, which can be difficult to achieve in hospital settings.

The need for reliable pupil assessment at various stages of clinical care (e.g. pre-hospital care, neuro-monitoring in critical care, etc.) and recent advances in smartphone cameras, have motivated efforts to develop a smartphone-based method to quantify the PLR. In addition, recently there has been a rapid development of neural network-based computer vision models ([11, 17])) that allow robust extraction of image features with a precision suitable for biomedical imaging, including subtle changes of the pupil size down to micron accuracy. In this context, a computational approach to lighting-invariant PLR measurements is a requirement to measure PLR reliably, across varying ambient light conditions.

In response to these challenges and needs, this study first aimed to establish the relation of each PLR parameter with lighting, and then develop lighting-invariant versions of the parameters, using an FDA-listed smartphone app to measure PLR. To correct for the effect of lighting on each pupil parameter, we used a two-step process. First, we fitted a model to predict the metric based on lighting, in healthy individuals. Second, this prediction was used to subtract away the effect of lighting. This leaves a lighting-corrected metric, that reflects how the metric deviates from what it should be, given the lighting. Second, we aimed to establish that these lighting-invariant parameters remain able to discriminate reactive and unreactive pupils. To this end, we used an anticholinergic agent tropicamide to dilate and render the pupil unreactive. Crucially, we developed the Pupil Reactivity (PuRe) score (PuRe), a scalar composite parameter which is based on the lighting-corrected PLR parameters and quantifies the pupil reactivity. Finally, we tested the performance of the lighting-invariant index in an unfavorable environment involving two distinct levels of ambient lighting.

## METHODS

### Laboratory calibration

First, using a smartphone-based pupillometer (AI Pupillometer, Solvemed Inc.) running on an iPhone 13 Pro, we acquired pupillometry data under various precisely controlled illumination conditions (Fig. 2A). We tested 9 healthy participants (male= 6, female= 3) in a quiet, dimly lit room. Each participant sat comfortably and placed their head on a chinrest, which was positioned 15 cm away from a phone camera mounted on a retort stand. We modulated the illumination in the room using a combination of adjustable LED lamps and dimmable light bulbs. We measured the illuminance using a digital luxometer, attaching its photometric head to the chinrest near the eye to be stimulated.

**FIG. 2:**
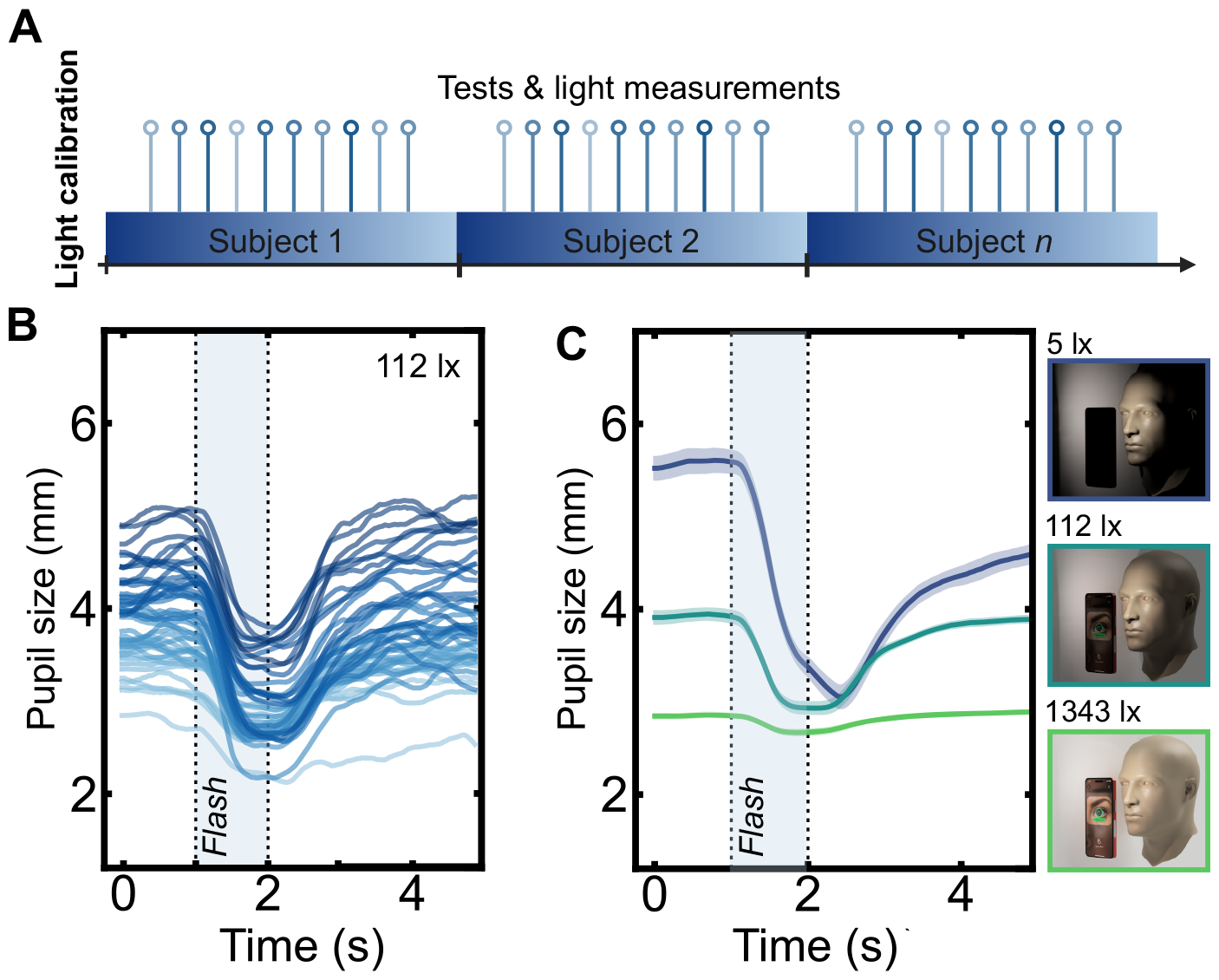
Raw pupillograms acquired under different conditions. **(A)** Design of the lighting study. Each participant was tested under a range of illuminations. **(B)** Raw inferred pupil size for each recording for the 9 participants, shown in the 112 lx condition. The pupil constricts in response to light. The flash time is depicted as a shaded window. **(C)** Mean traces in the lighting study. Shaded areas indicate the standard error of the mean. As expected, both the baseline pupil size and response amplitude were both smaller under brighter illumination.

We allowed each participant a minimum of 10 minutes to acclimatise to the respective lighting conditions before initiating the testing. At each illumination, five repeated measurements were taken, at 30 s intervals. We elicited the pupillary light reflex using a light pulse with a duration of 1,000 ms and an illuminance on the face of approximately 300 lx, generated by the iPhone’s LED flash. Video recordings were acquired at 60 Hz, with each recording five seconds long. The recording included one second prior to the light stimulus, one second during the stimulus, and three seconds post-stimulation.

### Clinical study at an outpatient ophthalmology clinic

In order to ensure lighting correction still enabled the detection of unreactive pupils, we rendered the pupil unreactive by administering tropicamide, an antimuscarinic drug commonly used to dilate pupils before the ophthalmological examination. *N* = 15 patients (female= 11, male= 4), mean age 55.1 ± 16.61 were recruited from the Oftalmika Eye Hospital. Ethical approval was granted by the Research Bioethics Committee at Nicolaus Copernicus University in Toruń, Poland (ref. KB 42712021). All patients provided informed written consent to participate in the study. The study complied with the International Conference on Harmonization Good Clinical Practice (ICH-GCP) guidelines. Patients were excluded if they had previous ocular surgery. The patients had a range of diagnoses from shortsightedness and far-sightedness to age-related cataracts and retinal breaks without detachments.

At the onset of each clinical pupillometry appointment, patients were seated in the clinic room under controlled lighting conditions. The study was performed in the dark, in routine ophthalmology clinic lighting conditions, ranging from 12 to 120 lux. The phone was held by the clinician, and the flash provided a backlight intensity of approximately 24 lx. The external lighting varied between approximately 11 to 120 lx as estimated by phone sensors (details below).

We administered a single drop of tropicamide 1% eye drops in each eye as part of the patients’ routine clinical care to induce mydriasis. We measured the pupillary light reflex at baseline, several minutes before administering the eye drops, in each eye. Reactivity was re-tested 15 minutes following administration, again in each eye, to evaluate the changes in pupil reactivity to light.

### Validation in an austere environment

During the NEP2NE (Nautical Experiments in Physiology, Technology and Underwater Exploration) scientific mission, 3 individuals performed various research tasks, one of which was to test whether hyperbaric saturation affected physiology at a depth of 22 feet, over 5 days. Three subjects (female= 1, male= 2; ages 35-55) self-selected to take part in a 120-hour scientific aquanautic mission in an underwater dive complex at a depth of 22 feet, and a pressure of 1.6 atm. Employing an AI-based smartphone software-as-medical-device (AI Pupillometer, Solvemed Inc), with no additional hardware, PLR was measured in each eye at 8 timepoints: at pre-exposure to the hyperbaric environment where the illumination was bright, and at seven scheduled intervals during the mission when illumination was dark. Ethical approval was provided by the University of Alberta Research Ethics Board (REB, Ref. Pro00131062).

The code and data used in this paper is publically available at https://github.com/solvemed/PuRe. As further data are added to the training set, this repository will be updated.

## PRE-PROCESSING

The phone camera captured video with a frame rate of 60 Hz. Each frame was passed through a custom convolutional neural network trained on a large set of manually-annotated images, to determine the absolute pupil size. Following this, we employed a proprietary signal processing pipeline to infer the pupil size over time. The inferred pupil size was then fitted to a canonical pupil response function, operationalised as the sum of two logistic sigmoid functions, with physiological constraints. This yielded parameters for the pupillary response, in accordance with standard quantitative definitions (**Table I**).

**TABLE I:** Parameters utilized for quantitatively assessing the dynamics of pupil response to light stimulation with the calculation methodology and their physiological meaning.

| Parameter | Abbreviation | Quantitative Definition |
| --- | --- | --- |
| Baseline Pupil Diameter | INIT | Mean pupil diameter calculated over the initial 250 ms period. |
| Minimum Pupil Diameter | END | Median of the smallest pupil diameter recorded within 130 ms during the first 1500 ms post light stimulation. |
| Final Pupil Size | FIN | Asymptotic final size of the pupil, after re-dilation. |
| Constriction Amplitude | CAMP | Computed as the difference between the Baseline Pupil Diameter (INIT) and Minimum Pupil Diameter (END). |
| Maximum Constriction Velocity | MCV | Represented by the minimum value of the first derivative observed during the constriction phase. |
| Peak Dilation Velocity | PDV | Denoted by the maximum value of the first derivative obtained during the dilation phase. |
| Constriction Latency | LAT | Interval between the onset of light stimulation and the commencement of pupil constriction. |
| Recovery Time to 75% Amplitude | T75 | Time for the pupil to recover 75% of the Constriction Amplitude, after the pupil reaches its minimum size. |

### Analysis and statistics

#### Effect of lighting

To determine whether lighting influences a given pupil parameter, a general linear mixed effects model was used to predict the parameter as a function of external lighting. The illumination was treated categorically to avoid assuming linearity, and a random intercept per subject was included to control for inter-individual differences, yielding the following model:

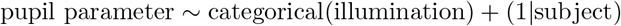

Analysis of variance was then used to test for significance. Linear models were fitted using the statsmodels package in Python.

#### Correction for lighting

Each pupil parameter was then corrected for lighting by first fitting a model to predict the pupil parameter from lighting and baseline pupil size (INIT parameter). This prediction was then used to re-scale the parameter.

Predictors of the pupil parameters included the camera exposure (Exposure, determined by the phone firmware), and the baseline pupil size (Baseline). Baseline pupil size is physiologically determined, but in healthy people does accurately track ambient lighting. Correction curves were fitted for each of the parameters in **Table I**, except for INIT which was used as a predictor. Each parameter was predicted by regression against linear and nonlinear terms of baseline pupil size and Exposure. The nonlinear terms were *X*^−1^, *X*^2^, *X*^2^, log *X*, log *X*^2^ where *X* can be Baseline or Exposure, as well as interactions Baseline × Exposure and log(Baseline) × log(Exposure). We used stepwise regression to optimise the Bayes information criterion (BIC) for each parameter. The top 250 models for each parameter were then fine-tuned using LASSO regularization (*α* = 0.006) yielding one winning model for each parameter.

We reasoned that a “corrected parameter value” should be proportional to the measured value, but be scaled in such a way that the value is the same across the range of lighting conditions. In order to perform this type of correction, we obtained a predicted value of the parameter for the given lighting, using the best fitting-model determined above. The corrected values were then computed as the deviation of the measurement from its lighting-predicted value, added to the parameter’s average value. For this, we used the mean values across all lighting conditions.

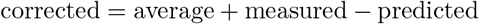

The motivation behind this formula is as follows. A person who has an exactly average pupil response (in this case, the average of the participants in this study), will have metrics equal to the predicted values. The formula will then yield the population average, plus zero. When the lighting changes, the prediction and metric will change in tandem, giving the same result irrespective of lighting. A person with different pupillary metrics (e.g. slower, faster, smaller in amplitude) will deviate from the prediction by a given amount. The formula assumes that this will be present at all given lighting levels, and yields the population average plus or minus this deviation. This produces a lighting-invariant index that remains sensitive to deviations of a person’s pupil responses from the normative value.

The methods used for computing the lighting-invariant index are general enough to be used on any model of phone. The specific coefficients presented in the results were all collected on a single model, and therefore apply to the iPhone 13 only.

## RESULTS

### Lighting influences all pupillometric parameters

First, we aimed to determine, under laboratory conditions, how PLR parameters were affected by varying the ambient illumination. The pupillograms, i.e. inferred pupil size over time time, obtained under 112 lx illumination are shown in Fig. 2B. Mean pupillograms differed greatly according to lighting, as predicted (Fig. 2C for three example lighting conditions: 5 lx, 112 lx, and 1343 lx). The eight parameters were calculated for each pupillogram. To quantify the dependence of each pupil parameter on lighting, the mean and the standard error of the parameter value across participants were computed (Fig. 3A-H).

**FIG. 3:**
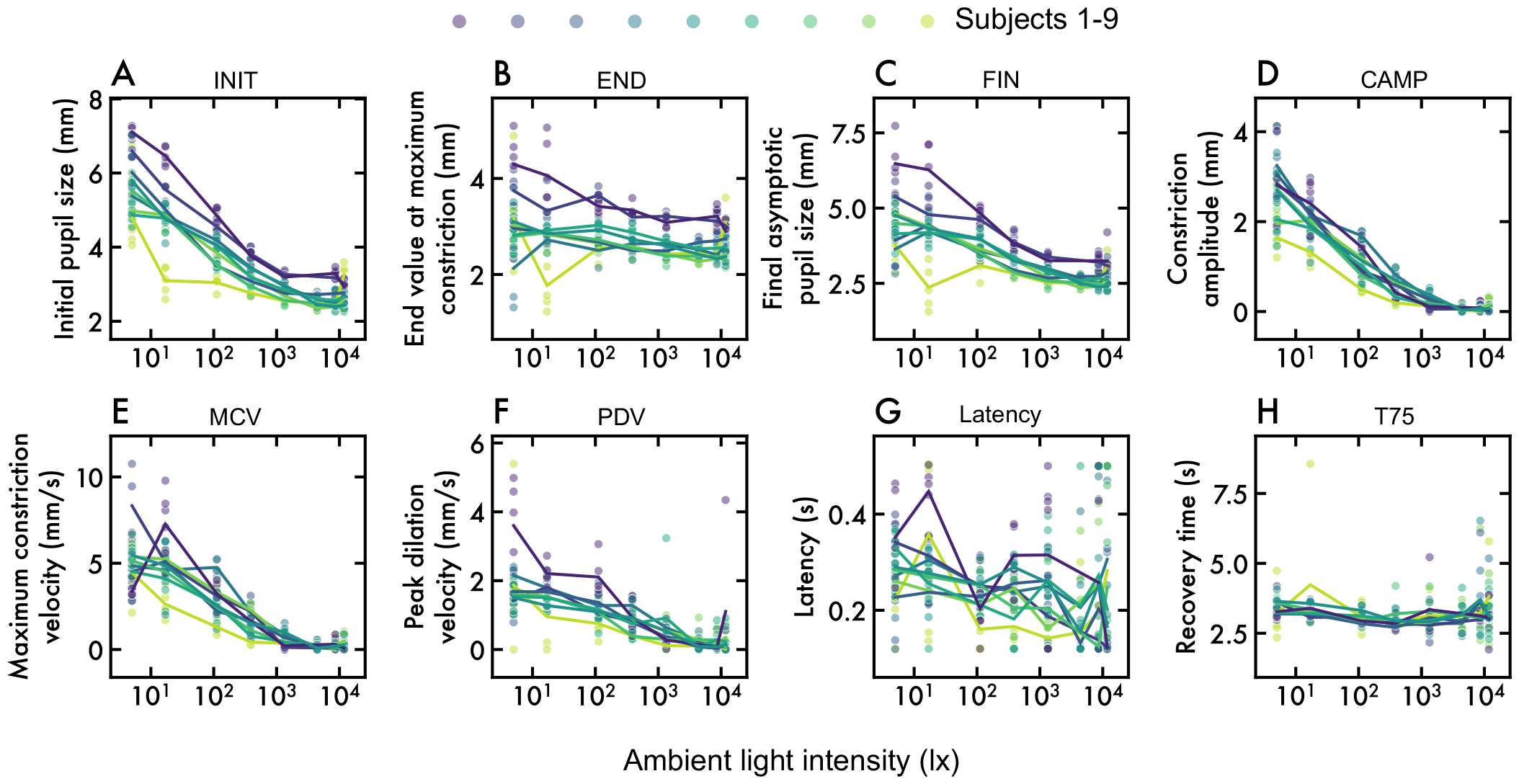
Impact of lighting on pupillometric parameters. **(A-H)**. The figures show the significant influence of lighting conditions on all evaluated pupillometric parameters. Each subject is in a different colour, with the line denoting the mean. **(A)** Baseline pupil diameter (INIT), **(B)** Constriction amplitude (CAMP), **(C)** Final asymptotic pupil size (FIN), **(D)** Constriction amplitude (CAMP), **(E)** Maximum constriction velocity (MCV), **(F)** Peak dilation velocity (PDV), **(G)** Latency (LAT), **(H)** Recovery time (T75).

Each parameter was strongly determined by lighting (general linear mixed effects model of pupil parameter vs external lighting with a per-subject random intercept and slope, all *p* < 0.05, all *F* (7, 338) > 4.45, **Supplementary Table I**). The most strongly affected parameters were baseline (INIT), constriction amplitude (CAMP), and maximum constriction velocity (MCV) (all *F* > 190). Specifically, brighter illumination decreased initial size, reduced constriction amplitude and slowed the maximum constriction velocity.

### Correction of each PLR parameter for lighting

Next, we sought to compute a “lighting-corrected” form of each PLR parameter. To do this, we examined the predictors of each parameter across lighting conditions (4A), which were used to compute corrected versions of each parameter (4B). Uncorrected and corrected values across the range of lighting were compared (Figure 4C). In each case, the predicted values were computed using leave-one-out cross-validation, so the model had not seen the data that was being corrected. The winning models are shown in **Table II**. To measure how well the correction works, we tested whether the parameter (before and after correction) varied with the lighting level. This yielded an *F* statistic for the uncorrected and corrected parameter. The light-dependence was reduced for each of the parameters.

**TABLE II:** Best model to explain the lighting-dependence of each of the standard PLR parameters. Each pupil parameter was predicted by a linear combination of the variables shown. The *R*^2^ statistic indicates the quality of the fit. The F statistic indicates how much each parameter varies with lighting, calculated for raw parameters and after applying the lighting correction.

| Pupil parameter | Ingredients of winning model | F-statistic before correction | F-statistic after correction | $R^2$ |
| --- | --- | --- | --- | --- |
| END<sub>c</sub> | Baseline + Baseline<sup>2</sup> + Exposure<sup>2</sup><br>+ log(Baseline) × log(Exposure) | 5.57 | 3.16 | 0.68 |
| FIN<sub>c</sub> | log(Baseline) + Baseline<sup>2</sup> + Exposure<sup>2</sup> | 63.83 | 5.58 | 0.87 |
| CAMP<sub>c</sub> | Baseline + Baseline<sup>2</sup> + Exposure<sup>2</sup><br>+ log(Baseline) × log(Exposure) | 268.93 | 2.71 | 0.90 |
| MCV<sub>c</sub> | Baseline + log(Exposure) + Baseline<sup>2</sup><br>+ log(Baseline) × log(Exposure) | 207.81 | 1.51 | 0.83 |
| PDV<sub>c</sub> | Exposure<sup>2</sup> + Exposure<sup>-1</sup> + log(Baseline) × log(Exposure) | 65.61 | 1.44 | 0.55 |
| LAT<sub>c</sub> | Baseline × Exposure | 6.60 | 1.26 | 0.07 |
| T75<sub>c</sub> | Exposure + log(Exposure) | 4.5 | 0.56 | 0.07 |

**FIG. 4:**
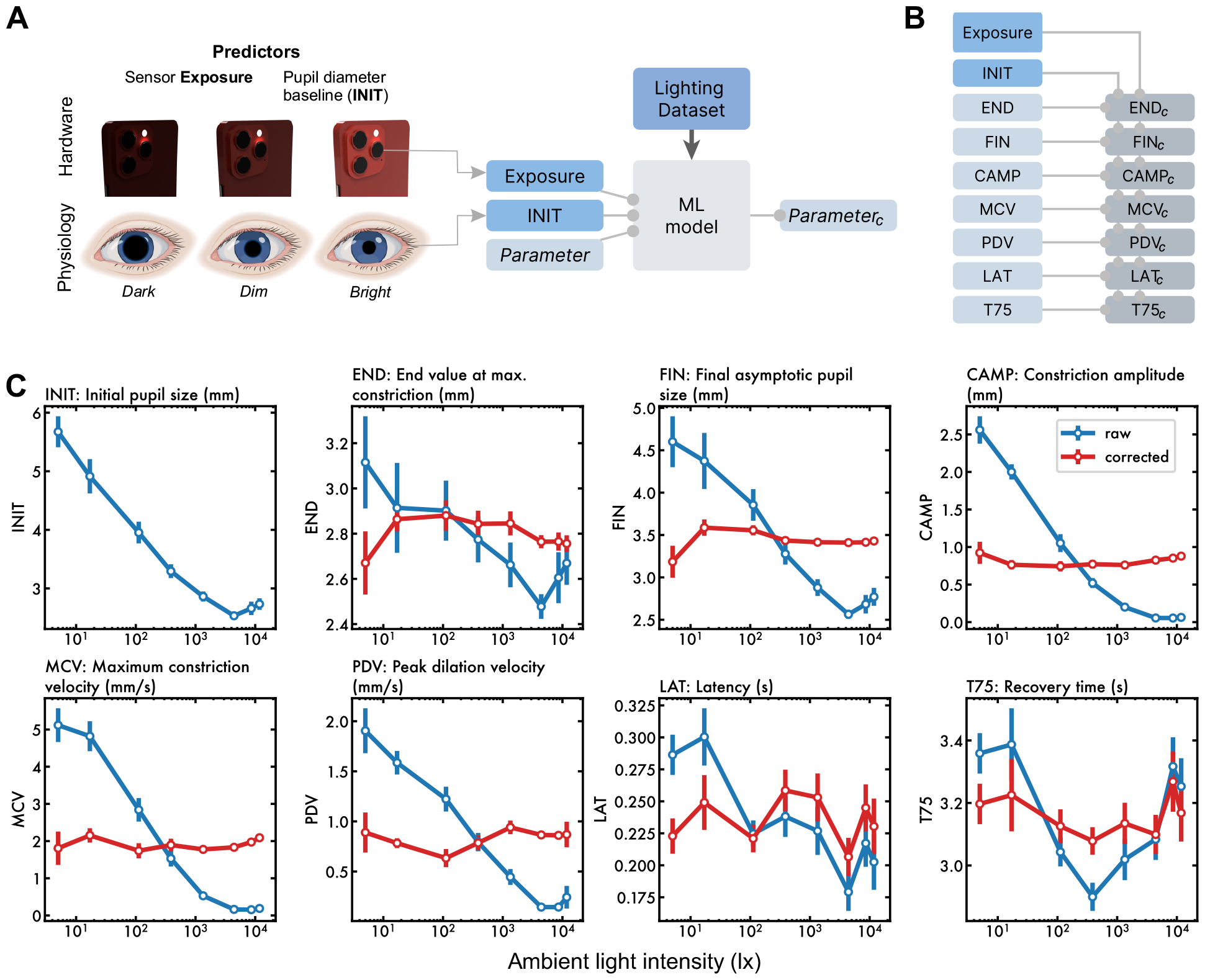
Lighting-corrected parameters. **A)** To predict how each parameter varies with lighting, hardware exposure measurements and the baseline pupil size are used as inputs to the model. The correction aims to make each measure lighting-invariant. (**B**) Each of the seven parameters is corrected using the trained models. The optimal model for each parameter is given in **Table II** (**C**) Each raw parameter was strongly dependent on illumination (blue). The lighting-corrected parameters (red) each showed a flatter dependence on illumination.

### Corrected PLR parameters remain able to detect unreactive pupils

It is possible that by enforcing a parameter to be insensitive to lighting, it may become insensitive to other important measures including pathology. We therefore asked how each of the corrected parameters fared at detecting unreactive pupils. This was tested by measuring PLR before and after tropicamide eye drops. The demographics of the patients are in **Table III**.

**TABLE III:** Demographics of participants in the tropicamide study.

|  |  |
| --- | --- |
| Number of participants | 15 |
| Age | $53.8 \pm \text{SD } 19.4$ |
| Sex | female = 11, male = 4 |

We compared the constriction parameters (END, FIN, CAMP, MCV, PDV, LAT, T75) before vs. 15 minutes after administration of tropicamide eye drops. The most accurate parameter for distinguishing drug condition was CAMP (Fig. 5A). After correction for lighting, several parameters distinguished clearly between pre- and post-tropicamide (Fig. 5B). Notably, the corrected CAMP parameter was 100% accurate with a wide margin of separation. As expected, the final pupil size did not discriminate well between tropicamide conditions, as it is highly correlated with the baseline pupil size and therefore becomes relatively invariant. A receiver operating characteristic was computed for each parameter, to distinguish pre vs post-tropicamide, using a kernel density estimate. The area under the curve for several parameters (END, CAMP, PDV, LAT, T75) was numerically greater after correction, compared to before correction (Fig.5C).

**FIG. 5:**
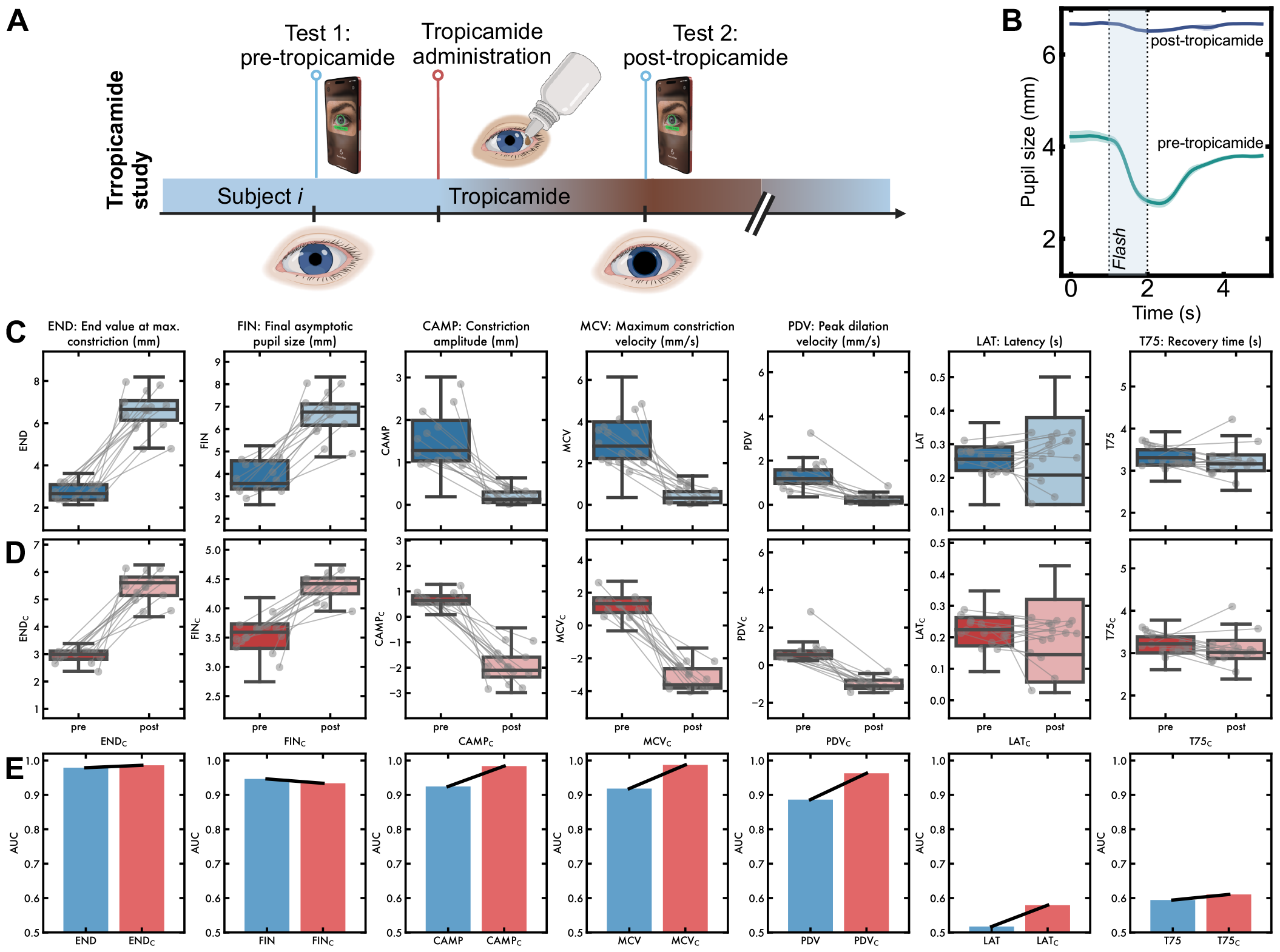
Effect of correction on PLR parameters under the effects of the mydriatic agent tropicamide. **(A)** Study design: participants were tested before and 15 min after tropicamide instillation. **(B)** Pupillary response to light after applying tropicamide drops, which leads to a fixed dilated pupil, giving an “artificial lesion”. **(C)** Raw values of parameters showed differences between pre- and post-tropicamide recordings but also showed considerable variability. **(D)** After lighting-correction, most parameters continued to discriminate strongly between pre- and post-tropicamide recordings. **(E)** Area under the curve for distinguishing between pre-tropicamide and post-tropicamide, calculated separately for uncorrected and lighting-corrected parameters. Discrimination improved numerically for all parameters except for final pupil size (FIN).

**FIG. 6:**
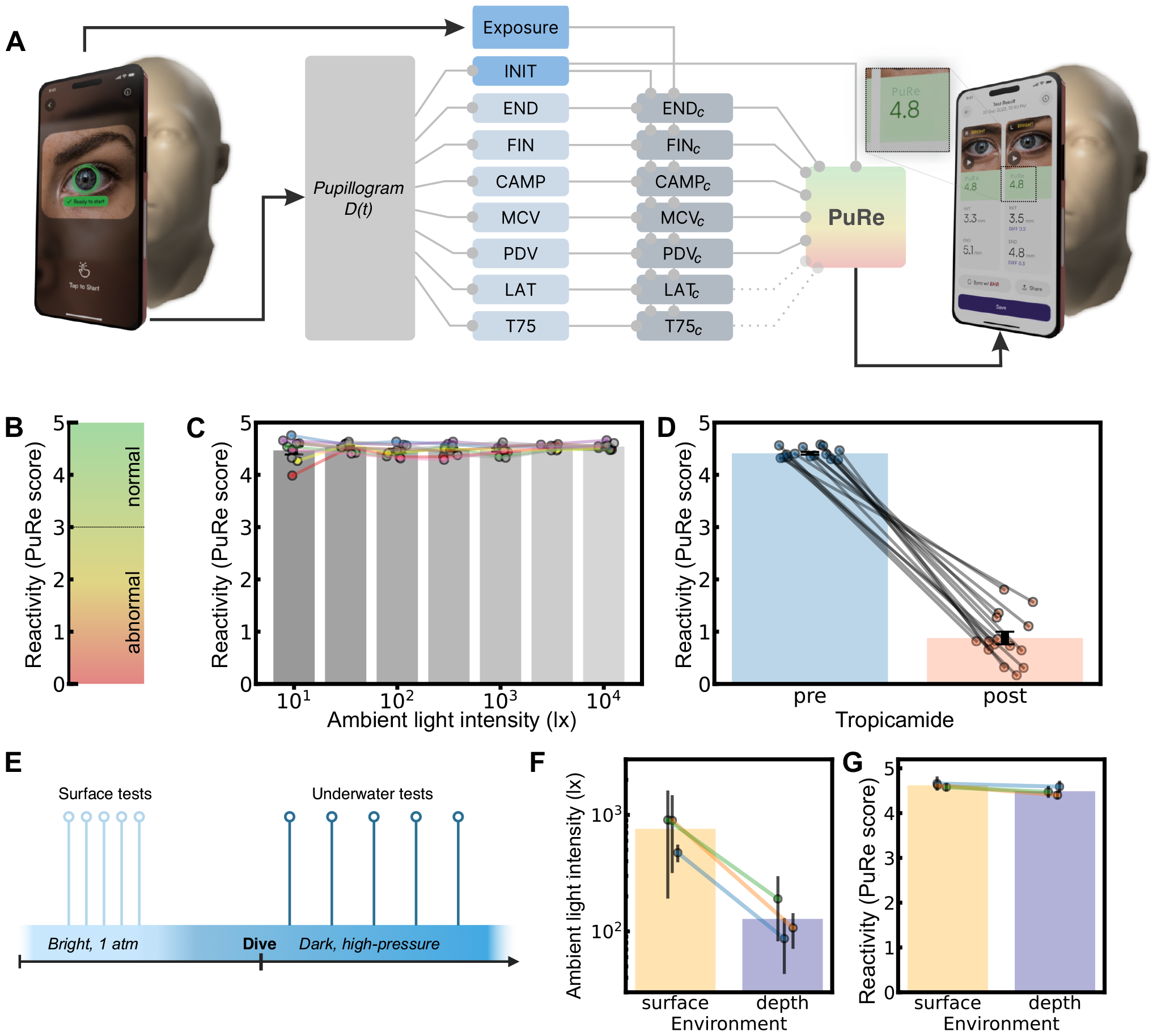
lighting-invariant index of pupil reactivity. **(A)** To calculate a score, pupil diameter *D*(*t*) was recorded and uncorrected parameters were extracted. Exposure and INIT were used to compute lighting-invariant parameters (see section). The parameters were combined to optimise discrimination between the pre- and post-tropicamide recordings, resulting in the Pupil Reactivity (PuRe) score PuRe. **(B)** the score was scaled to lie in the range 0-5, based on the distribution of normative values. A value of 3.0 corresponds to the midpoint between reactive and unreactive pupils. **(C)** The Pupil Reactivity (PuRe) score PuRe is plotted in the lighting study. Bars are shown as a function of varying ambient illumination, demonstrating relative invariance to lighting. **(D)** To demonstrate that the score still distinguishes well between reactive and non-reactive pupils, the score (PuRe) is shown before tropicamide drops, and 15 minutes after. the score maintained a good separation of the two conditions. **(E)** In the NEP2NE study, three healthy subjects were tested on the surface and after diving into an underwater habitat. **(F)** Lighting was significantly brighter at the surface compared to depth conditions. **(G)** The Pupil Reactivity (PuRe) score was unaffected by the conditions.

### Optimising discrimination of unreactive pupils using a score that combines corrected pupil parameters

Although individual corrected parameters already provide a strong separation of tropicamide from control, including multiple parameters enables the detection of more subtle changes including slowing of the PLR, changes in amplitude, and the ability to re-dilate. Therefore we estimated the optimal hyperplane to separate the reactive and unreactive pupils using regularised multi-variable logistic regression. We included the seven corrected parameters, and their powers of 2 and -1, and their first-order products. LASSO regularisation was applied using a coefficient of *α* = 0.006, and included a penalty in the regression for light-dependence to maintain light invariance.

The logistic regression yielded coefficients which provide a score of reactivity with a formula (three decimal places):

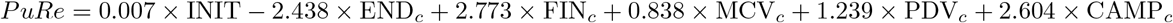

In order to correspond to commonly used clinical scales, we scaled our Pupil Reactivity (PuRe) score using a piecewise function to the range 0-5 (0, non-reactive pupil; 0-3, abnormal/”sluggish” response; 3-5, normal/brisk response). The line segments making up the function are calculated to map specific parts of the distribution of scores onto specific points on the final scale. The function maps median(post) − 3 × stdev(post) ⟶ −0.5, median(post) − stdev(post) ⟶ 0.2, median(post) ⟶ 0.8, median(post) +stdev(post) ⟶ 1.0, 0 ⟶ 3, median(pre) − 3 × stdev(pre) ⟶ 4.0, median(pre) ⟶ 4.5 and where median, stdev denotes median and standard deviation of values of PuRe obtained before mapping, before tropicamide administration (pre) and after tropicamide administration (post).

### Validating the lighting-invariance of the reactivity score during the NEP2NE underwater study

Finally, we aimed to test the lighting-invariance of the reactivity score in an environment less favourable than the lab or clinic. The AI Pupillometer app was deployed in a hyperbaric underwater lagoon, and tests were performed at the surface and at depth. The illumination measured by the sensors was over six times brighter at the surface, 790 ± 570 lx (mean ± SD), than at depth, 128 ± 83 lx (linear mixed effects model with the environment as predictor, and subject as group variable, random intercept, *t*(environment) = 57.5, *P* > |*t*|< 0.001). There was no difference in the reactivity index (PuRe) comparing surface vs depth (*t*(environment) = 2.93).

## DISCUSSION

Despite its crucial role in emergency and critical care, the pupillary light reflex (PLR) is highly dependent on background illumination. We quantify and fit the relationship between each PLR parameter and illumination, allowing us to develop lighting-invariant versions of each parameter. Crucially, we demonstrated that they retain sensitivity to discriminate key pathological features of an unreactive pupil. We then combined these lighting-invariant parameters to produce a single reactivity index (PuRe) that optimally discriminated unreactive pupils. Since the score is based on these corrected parameters, it is tuned to be stable under a wide range of lighting conditions. The lighting-invariant parameters described here are a step towards increasing the reliability and accessibility of PLR measures. This is especially important as conventional infra-red pupillometer devices struggle with varying ambient light (REF Ong, C. 2019), which can lead to inaccurate readings and sub-optimal clinical decision-making.

Quantitative pupillometry plays an increasingly important in critically ill patients, both for the evaluation of neurological status ([14, 16, 28]), monitoring ([16, 28]), and prognostication ([16, 26, 28]). These especially include patients affected by acute brain injuries ([4, 26]), where there is a need to detect deterioration of neurological state. Quantitative pupil dynamics have also gathered interest in recent years in neurology ([8]), neurosurgery ([4, 7]), anesthesiology ([27]), and emergency medicine ([3]). Objective and accurate pupil size and reactivity testing have been shown to be clinically valuable, reducing assessment errors and the examiner’s uncertainty, and aiding nurses and clinicians in improved clinical decision-making compared to when a subjective visual assessment of pupils is performed ([22, 23]).

Even when measured with infra-red pupillometer devices, PLR parameters can vary with lighting. In one study of critically ill patients, pupil reactivity differed significantly under different lighting conditions ([24]). Standardizing lighting conditions is therefore necessary to maximize measurement reliability, but during routine clinical care, this is extremely challenging, for many practical reasons. Using opaque eye-cups (rubber cups) to block ambient light during the measurement has been suggested as a possible solution ([10]), however, this is not possible with most pupillometer devices. Further, it may not fully alleviate ambient light effects in conscious patients due to the consensual pupillary response and adds costs. Thus, robust lighting-correction methods are needed to allow for accurate and clinically meaningful pupil reactivity tracking in patients.

Naively, one might think that the change in pupil size, as a proportion of the baseline size, might correct for background effects, but this is not the case ([13], [30]). In our data, the lighting dependence of various parameters was nonlinear, and optimal correction differed according to the parameter. Furthermore, the optimal axis for discriminating reactivity required a non-intuitive combination of parameters, though constriction amplitude was the most dominant, as expected.

The final underwater study demonstrated invariance not only to illumination, but also to hyperbaric conditions. While the effects of hypobaric conditions on brain function are well established ([33]), such as those at high altitudes with diminished P_*i*_CO_2_ levels, the effects of hyperbaric pressures remain poorly understood. Saturation diving complexes offer several challenges, owing to their isolation, confinement, and location in hyperbaric environments. Even after several days at depth, at 1.6 atm, we found no effects of pressure on the pupillary reactivity index after correcting for lighting. The findings underscore the stability of PLR under different lighting situations and in a hyperbaric environment. Moreover, we observed that smartphone-based pupillometry was easy to deploy in the austere environment owing to its lightweight, portable form factor. This last study was opportunistic. While the results showcase the robust mobile platform capable of remote operation with local processing, further tests in a real intensive care setting are needed.

While the numerical results presented were calculated using only one model of the phone, the same calibration can be applied to data from other models. Since different phones have different cameras with different resolutions, sensitivities, and exposure relationships, calibrating individual phones is likely to be essential to producing reproducible results. The methods proposed in this paper provide a framework for how this should be done.

We argue that lighting-invariant quantitative pupillometry could herald a new area for pupillometry. First, lighting-corrected parameters may be more clinically useful compared to their raw counterparts, since they are more robust to environmental conditions and may offer more sensitive discrimination between reactive vs “sluggish” vs unreactive pupils. Lighting-corrected parameters are also likely to improve the quality of pupil reactivity trends, crucial for detecting neurological deterioration, for example in critical care settings. Presently, gradual changes in ambient light (for example as a consequence of typical changes in sunlight over a diurnal cycle) can induce false trends in pupil reactivity and lead to sub-optimal clinical decisions. Second, it allows reliable pupil assessment across the spectrum of care. Much of the data in this study was collected from patients by clinicians in a clinical setting. Adopting the lighting-corrected open-sourced Pupil Reactivity (PuRe) score could be an important step towards the standardization of pupil testing across different healthcare professionals and research groups. Since it can be performed using a standard smartphone app with no additional hardware, quantitative pupillometry opens new opportunities to measure neurological vital signs, that have historically required a healthcare professional. For example, with the ability to perform rapid quantitative pupil assessment reliably in a variety of environments, emergency medicine technicians may be able to better triage brain injury patients at the scene of accidents and neuro-monitor the patients during transport, with the potential for improved decision-making, greater cost-effectiveness, and improved outcomes. In addition, measurements captured with a smartphone can be shared flexibly across clinical teams providing intensive care and surgical teams, providing richer baseline data and thus facilitating clinical assessment, prognostication, and decision-making.

Finally, the Pupil Reactivity (PuRe) score, presented here for the first time, represents the first-ever lighting-corrected score quantifying pupil reactivity. The score is directly applicable to clinical practice allowing for reliable and convenient assessment and tracking of pupil reactivity, necessary for evaluation and monitoring of patient’s neurological status. Importantly, the formula behind the score was presented here openly for the benefit of the clinical research community.

## Data Availability

The data will be made available by the authors, by request to the corresponding author.

